# Genetic Diversity of the *cagA* gene of *Helicobacter pylori* strains from Sudanese Patients with Different Gastroduodenal Diseases

**DOI:** 10.1101/19007435

**Authors:** Hadeel Gassim Hassan, Abeer Babiker Idris, Mohamed A. Hassan, Hisham N. Altayb, Kyakonye Yasin, Nazar Beirage, Muzamil M. Abdel Hamid

## Abstract

**Background:** There is an increase in the prevalence of *Helicobacter pylori* infection in Sudan, accompanied by a high incidence of upper gastrointestinal malignancy. The cytotoxin-associated gene *cagA* gene is a marker of a pathogenicity island (PAI) in *H. pylori* and plays a crucial role in determining the clinical outcome of *Helicobacter* infections.

**Objective:** This study aimed to determine the frequency and heterogeneity of the *cagA* gene of *H. pylori* and correlate the presence of *cagA* gene with clinical outcomes.

**Materials and methods:** Fifty endoscopy biopsies were collected from Fedail and Soba hospitals in Khartoum state. DNA was extracted using the Guanidine chloride method followed by PCR to amplify *16S rRNA* and *cagA* gene of *H. pylori* using specific primers. DNA amplicons of *cagA* gene were purified and sequenced. Bioinformatics and statistical analysis were done to characterize and to test the association between *cagA* gene and gastric complications.

**Results:** *CagA* gene was detected in 20/37(54%) of the samples that were found positive for *H. pylori*. There was no association between endoscopy finding and the presence of the *cagA* gene (p = 0.225). Specific amino acid variations were found at seven loci related to strains from a patient with duodenitis, gastric ulcer, and gastric atrophy (R448H, T457K, S460L, IT463-464VA, D470E, A482Q, KNV490-491-492TKT) while mutations in cancerous strain were A439P, T457P, and H500Y.

**Conclusion:** Disease-specific variations of *cagA* of *H. pylori* strains, in the region of amino acid residues 428-510, were evident among Sudanese patients with different gastroduodenal diseases. A novel mutation (K458N) was detected in a patient with duodenitis, which affects the positive electrostatic surface of *cagA*. Phylogenetic analysis showed a high level of diversity of *cagA* from Sudanese *H. pylori* strains.

## 1. Introduction

*Helicobacter pylori (H. pylori)* a bacterium is responsible for various gastrointestinal diseases, including gastritis, peptic ulcers, frequent bleeding events, gastric mucosa-associated lymphoid tissue (MALT) lymphoma, and carcinoma. ^(1, 2)^. A majority of infected individuals with *H. pylori* are asymptomatic^(3, 4)^and despite treatment, some infected patients may not clear the infection, thus leading to chronic complication. The possible reason for the various outcomes of *H. pylori* infection may relate to the variations in virulence factors of *H. pylori* strains in addition to host, environment, and dietary factors. ^(5)^

The cytotoxic associated gene A (*cagA*) is one of the most virulence factors which has been implicated as a possible marker to distinguish variations in the virulence of H. pylori strains. ^(6, 7)^The *cagA* gene encodes a 120-145kDa protein, and it is located within the ∼40kb pathogenicity island (*cag* PAIs). ^(5)^ The *cag* PAIs expresses a type IV secretion systems (T4SS) which introduces the cagA effector protein into the host target cells where it undergoes tyrosine phosphorylation by Src and Ab1 kinases^(8)^. The *cagA* protein is actively involved in the regulation of the spreading, migration, and adhesion of cells known to play a decisive role in mutagenic signal transduction, and stimulation of phosphatase activity also it induces vacuolation of primary human mucosal epithelial cells. ^(9, 10)^ *CagA* is the most intensely studied since its first association with the risk of gastric cancer. ^(10)^ Various studies have documented that infection with *H. pylori* strains expressing *cagA* was correlated with the development of severe gastrointestinal diseases such as atrophic gastritis, peptic ulcer, and gastric carcinoma. ^(11-15)^

*H. pylori* infect about half of the world population, but only a subset of 1% to 2% of those infected individuals will develop gastric malignancies. ^(1)^ The prevalence in developed countries is low 25%, a contrast to developing countries where the incidence is very high up to 90% ^(4, 16, 17)^ and the infection occurring early in life and tending to persist for longtime unless treated. ^(18)^There is an increased in the prevalence of *H. pylori* infection in Sudan accompanied with high incidence of upper gastrointestinal malignancy (13.2%) when compared to previous study reported by Elhadi *et al*.^(19)^ Therefore, the aim of this study was to determine the frequency and heterogeneity of *cagA* and its correlation with severe gastroduodenal diseases among Sudanese patients. To our knowledge, this is the first study in Sudan that reports the association of *cagA* and its heterogeneity to clinical outcomes of *H. pylori* infection. The molecular characterization of *cagA* and its mutations in amino acid sequence 428-510 were analyzed by using *in silico* tools. According to the X-ray crystallographic analysis of the N-terminal *cagA* fragment (residues 1-876) of the reference strain (*H. pylori* strain 26695),^(20)^ amino acid sequence 428-510is located in domain II which tethers CagA to the plasma membrane by interacting with membrane phosphatidylserine. It is characterized by a sizeable antiparallel β-sheet and an insertion of a subdomain (residues 370-446) between β5 and β8 strands. ^(21)^

## 2. Materials and methods

### 2.1 Study design and study population

A prospective cross -sectional hospital-based study was conducted in Fedial and Soba hospitals in Khartoum state between February and May 2016. Patients who referred to hospitals for upper gastrointestinal tract endoscopy and suspected to be infected with *H. pylori* were enrolled. Patients who had received nonsteroidal anti-inflammatory drugs and proton pump inhibitor (PPI) were excluded from the study, and none of the patients had recently been prescribed antibiotics.

### 2.2 Clinical Samples

The ethical approval was granted by the Institute of Endemic Diseases. All study participants completed informed consent forms prior to the commencement in the study.

A total of fifty patients were selected for the study, twenty-five of these patients were males, and twenty-five were females. Specimens were obtained from all patients and tentatively diagnosed by the gastroenterologist as having a helicobacter infection. The samples were labeled and transferred to the laboratory for DNA analysis. Patient endoscopy findings included gastritis, gastric ulcer, atrophy gastric, gastritis, and duodenitis.

### 2.3 Extraction of DNA and PCR analysis

DNA was extracted using Guanidine chloride method ^17^. PCR amplification was done using specific primers for *H. pylori* 16S rRNA gene F:5’GCGACCTGCTGGAACATTAC-3’ and R:5’GCGTTAGCTGCATTACTGGAGA-3’. A ready master pre-mix (Maxime, iNtRON BIOTECHNOLOGY, Korea) was used. A total volume of 25µl contained a1μl of each primer (10 pmole),1μl of DNA template and 22μl of de-ionized sterile water. A PCR reaction was carried out using a thermocycler (SensoQuest, Germany) following a protocol described by William *et al*. ^(22)^*CagA* gene was amplified using primer set F:5’ATAATGCTAAATTAGACAACTTGAGCGA-3’ R:5’TTAGAATAATCAACAAACATCACGCCA-3’ using a method defined previously. ^(23)^ PCR amplicons were analysed on 1.5% Agarose gel stained with 1µl ethidium bromide (10mg/ml). Electrophoresis was done at 90V and 25mA. DNA bands were estimated using a 100bp DNA ladder (iNtRON BIOTECHNOLOGY, Korea European Biotech). The bands were visualized using Gel documentation systemBioDoc-It UVP, Cambridge UK). DNA purification and sequencing was performed commercially at Macrogen Inc, Korea.

### 2.4 Statistical and Bioinformatics analysis

Data were analyzed using IBM SPSS Statistical software (Statistical Package for the Social Sciences). Chi-square was used to test the association of gastric complications with *cagA* gene. The nucleotide sequences of the cagA gene were compared for genetic diversity using nucleotide BLAST (http://blast.ncbi.nlm.nih.gov/Blast.cgi). ^(24)^ from NCBI, Highly similar sequences were retrieved and subjected to multiple sequence alignment using Clustal Omega - EMBL-EBI. ^(25)^ For nucleotides translation, evolutionary analysis and building a phylogenetic tree, GeneMarks ^(26)^and jalview^(27)^ were used, respectively. Modeling of 3D structural Protein was done by sending the protein sequence of the reference strain (*H. pylori* strain 26695)^(20)^ to raptor X online server. ^(28)^ Structural homology modeling of this reference strain (NP_207343) was done due to the existence of short disordered stretches, common in all crystal forms,^(21, 29-31)^ which their electron densities are not observed in any crystal such as residues 479-488 that is located in our region (residues 425-510). The Predicted structure was analyzed by using UCSF Chimera (version 1,11,2). ^(32)^

The nucleotide sequences of the *cagA* genes were deposited in the GenBank database (National Center for Biotechnology Information; https://www.ncbi.nlm.nih.gov/), Accession number of the strains with their clinical outcomes are presented in Table 1.

**Table 1.** Shows the accession number of the stains with their clinical outcomes

| **Name of strain** | **Outcome** | **Accession number** |
| --- | --- | --- |
| **Sudan-1** | Normal finding | MK074988 |
| **Sudan-2** | Normal finding | MK074989 |
| **Sudan-3** | Normal finding | MK074990 |
| **Sudan-4** | Gastritis | KX879074 |
| **Sudan-5** | Gastritis | MK074991 |
| **Sudan-6** | Gastritis | MK074992 |
| **Sudan-7** | Gastritis | MK074993 |
| **Sudan-8** | Gastritis | KX879073 |
| **Sudan-9** | Duodenitis | MK074994 |
| **Sudan-10** | Duodenitis | KX879075 |
| **Sudan-11** | Cancer | MK074995 |
| **Sudan-12** | Gastric atrophy | MK074996 |
| **Sudan-13** | Gastric ulcer | MK074997 |
| **Sudan-14** | Gastric ulcer | MK074998 |
| **Sudan-15** | Gastric ulcer | MK074999 |
| **Sudan-16** | Gastric ulcer | KX879072 |

## 3. Results

### 3.1 Endoscopy finding

Endoscopic examination was performed on all patients (n=50). Abnormal endoscopic finding was observed in 42/50(84%) of the patients, out of this 18/50(36%) were diagnosed with gastritis, 3/50(6%) with duodenitis, 12/50(24%) with gastric ulcers, 2/50(4%) with atrophy gastric, and 7/50(14%) with gastritis duodenitis.

### 3.2 Positive biopsy for *H. pylori* (16S rRNA)

Fifty (n=50) samples were tested for the presence of 16S rRNA, 37/50(74%) were found to be positive, and 13/50(26%) were found to be negative for *H. pylori*. (Figure 1).

**Figure 1:**
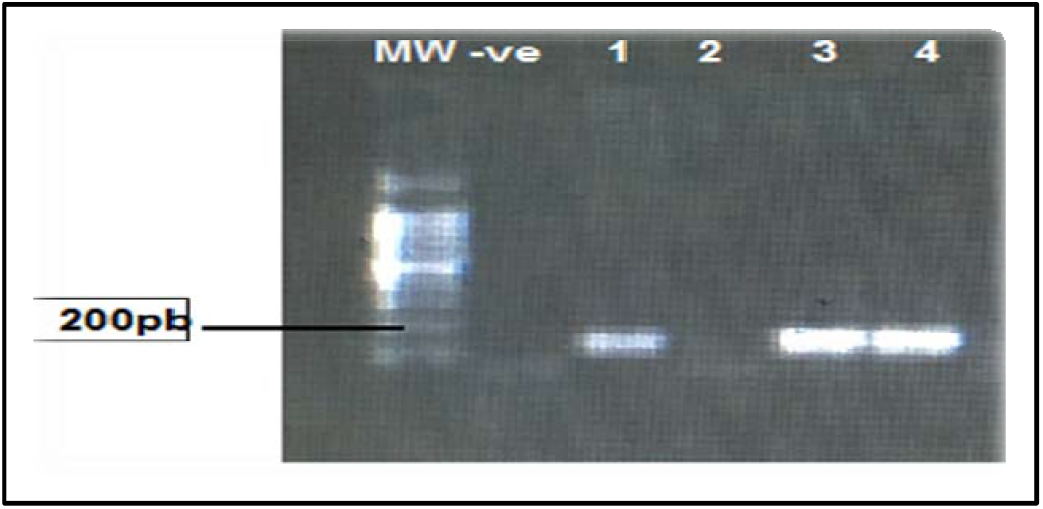
Amplified *H. pylori*16SrRNAgene PCR products from endoscopy biopsy. Lane 1: negative control (1), lanes (2,4,5) is16srRNAof *H*.*pylori* positive (168pb); MW: 100 bp DNA molecular weight marker

### 3.3 Frequency of *cagA* gene

The *cagA* gene frequency was found to be 20/37 (54%) among positive samples for (16S rRNA) of *H. pylori*. In gastric atrophy patients, it was found to be 2/20(10%),4/20(20%) in normal findings endoscopy patients,6/20(30%) in gastric ulcer patients,2/20(10%) in duodenitis patients, and gastritis patients were found to be 4/20(25%). (Table 2).

**Table 2.** Frequency of *cagA* gene in *H. pylori* strains from various gastroduodenal disease.

| **Endoscopic**<br>**finding** | **Endoscopy**<br>**finding frequency %**<br>**(N=50)** | **Number of specimen**<br>**(+ve)16SrRNA**<br>**(N=37)** | **Number of cag A**<br>**positive strains %**<br>**(N=20)** |
| --- | --- | --- | --- |
| **Normal** | 8 (16%) | 5(14%) | 4(20%) |
| **Duodenitis** | 3(6%) | 3(8%) | 2(10%) |
| **Ulcer** | 12(24%) | 10(27%) | 6(30%) |
| **Gastritis** | 18(36%) | 10(27%) | 4(20%) |
| **Gastroduodenitis** | 7(14%) | 6(16%) | 1(5%) |
| **Atrophy gastric** | 2(4%) | 2(5%) | 2(10%) |
| **Cancer** | 1(2%) | 1(3%) | 1(5%) |

### 3.4 Variations of amino acid among *cagA* protein

In comparison with strain 26695 (reference strain), disease-specific variations at different *cagA* loci were detected at a region in domain II (amino acid sequence 428-510) as illustrated (Figure2 and Figure 4).The mutations in amino acids R448H, T457K, S460L, IT463-464VA, D470E, A482Q, and KNV490-491-492TKT were found in *cagA* of duodenitis, gastric ulcer and gastric atrophy caused strains while mutations in cancerous strain were A439P, T457P, and H500Y. The amino acids at 467 and 497 loci were mutated in all strains regardless of gastrointestinal diseases.

**Figure 2:**
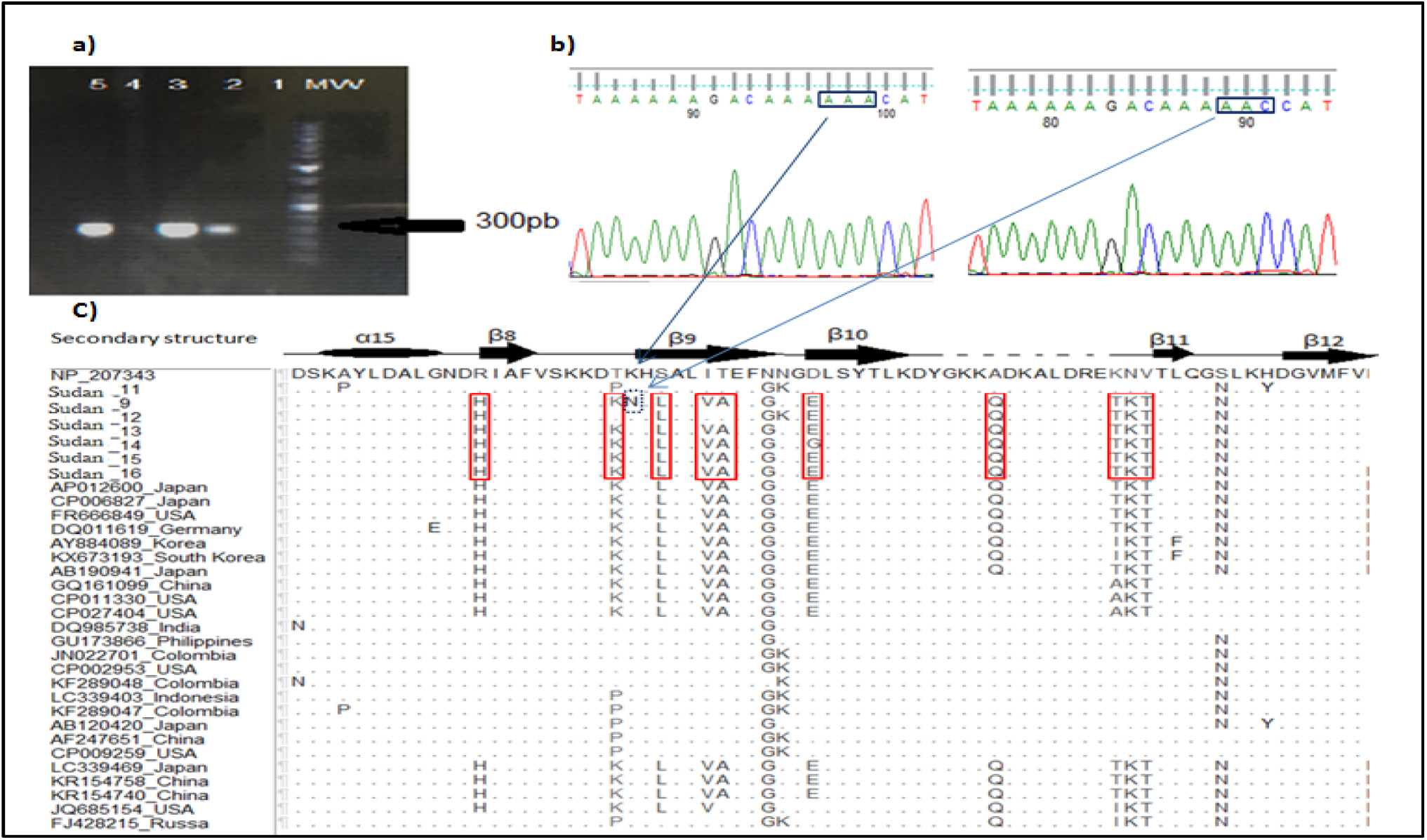
a) PCR gel showing bands corresponding to the *H. pylori cagA* gene from endoscopy biopsy. Lane 1: negative control (1), lanes 2,3,5 are Cag A positive genes of *H. pylori* (298pb), lane 4 negative sample.MW: 100bpDNA molecular weight marker. b) Shows the sequence chromatogram of codon 458 in normal finding and duodenitis causing strains, respectively by using Finch TV. c) Multiple sequence alignment of *cagA* gene sequences with other selected global strains obtained from GenBank. The different loci are shown in black letters, and white dots indicate similarity. The red boxes highlight amino acid variations at seven loci that are related to gastric ulcer, duodenitis, and gastric atrophy caused strains. Sudan-11 strain represents a cancerous strain. Secondary structure referred to strain 26695 ^(21)^.

**Figure 4:**
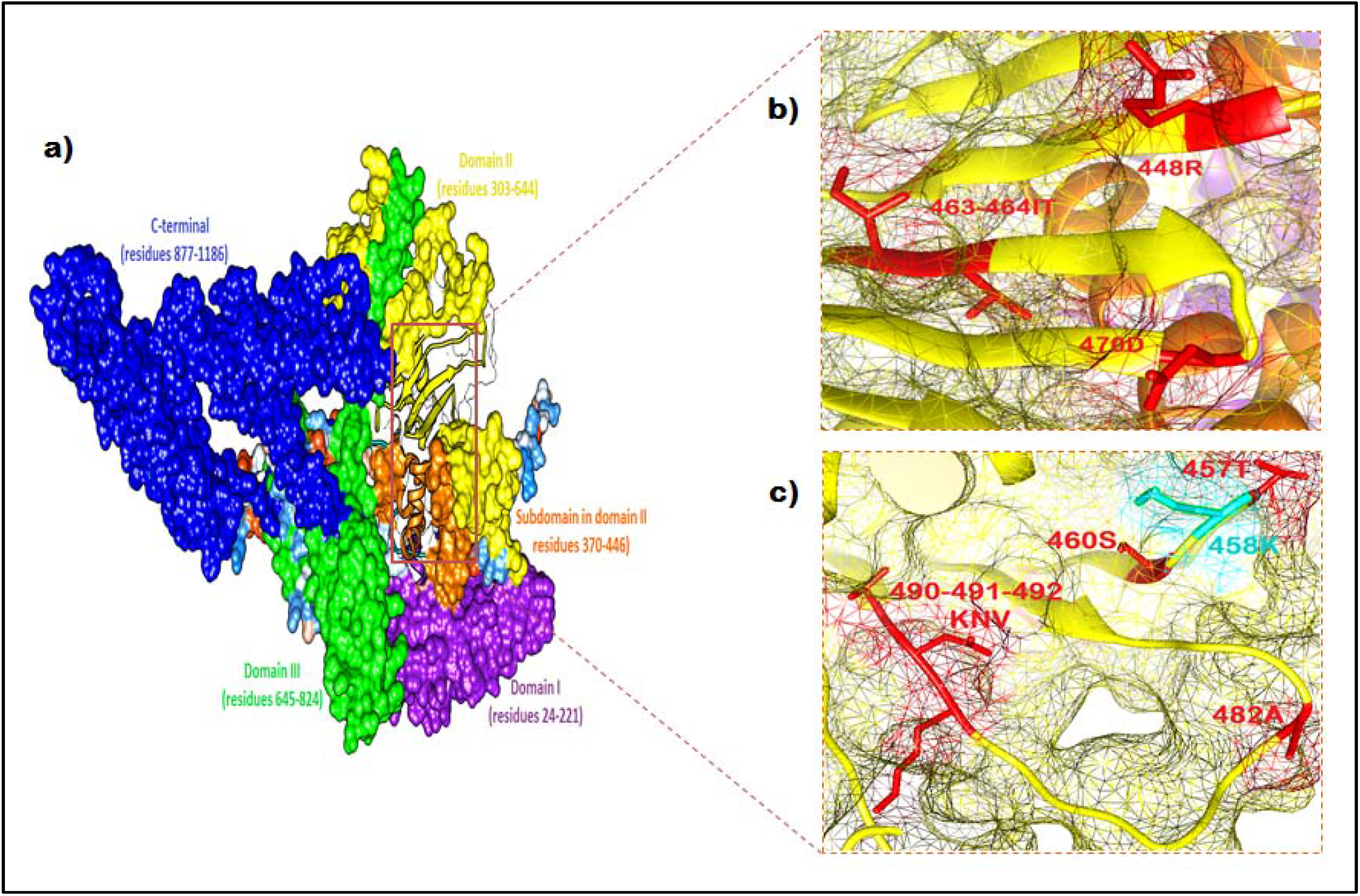
a) Structural homology modeling of *cagA* derived from *H. pylori* strain 26695^(20)^. b) and c) Shows loci of specific-disease mutations observed in duodenitis, ulcer, and atrophy gastric strains. The cyan color represents the locus of the significant novel amino acid substitution (458K) in duodenitis strain.

### 3.5 Phylogenetic analysis

Phylogenetic analysis of *cagA* with global *H. pylori* strains revealed that the Sudanese *H*.*pylori* strains were divided into two major groups. One group involves strains that cause duodenitis, gastric ulcer, and gastric atrophy while cancerous strain belongs to another common ancestor, as illustrated in Figure5.

**Figure 2.**
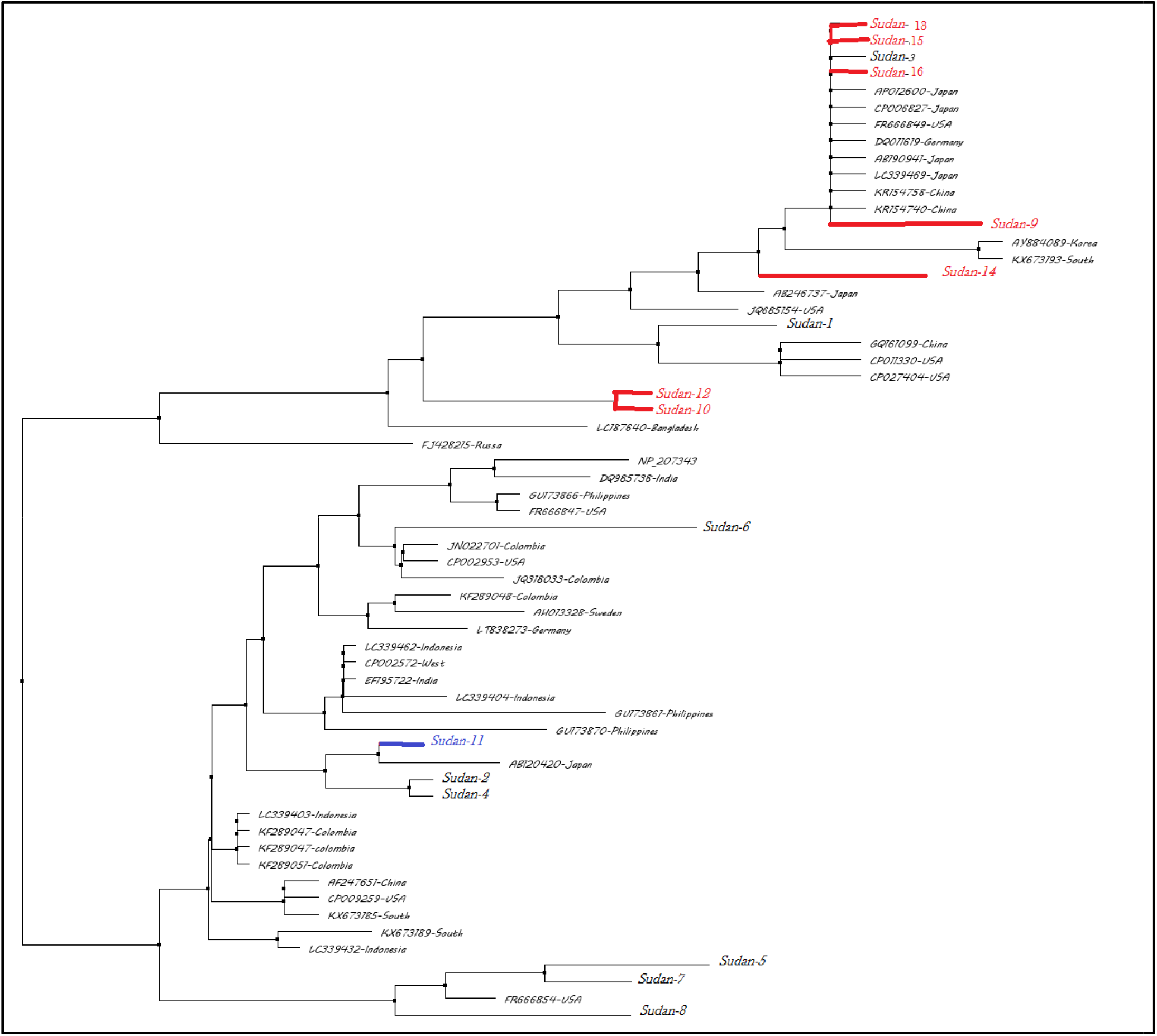
The neighbor-joining tree for the *Helicobacter pylori cagA* gene of 16 Sudanese strains compared with 20 global reference strains from the Gen Bank database. The Sudanese strains are divided into two major groups Strains that cause duodenitis, gastric ulcer and gastric atrophy are colored in red while the blue color represents cancerous strain.

## 4. Discussion

In this study, the analysis of amino acid sequence 425-510 of *cagA* indicated that the mutations at this region are distinguishable between gastrointestinal diseases. This indication is in agreement with the result obtained by Sung Soo *et al*., *2015* which suggested that the mutations of *cagA* of *H. pylori*, especially in C-terminus, are distinguishable between duodenal ulcer and gastric carcinoma. ^(33)^ In Sudan-9 strain that caused duodenitis, a novel mutation in lysine residue at 458 which constitutes with other lysine residues located in β9, β10 and α18; and two arginine residues (R624 and R626) the basic amino-acid cluster. ^(21)^ The basic amino-acid cluster provides a positive electrostatic surface potential, which is crucial for the CagA and phospholipid of the plasma membrane interaction. This interaction is essential for the delivery of CagA into host cells and also for the localization of delivered CagA to the inner face of the plasma membrane in epithelial cells. ^(34)^

Specific amino acid variations were found at seven loci (R448H, T457K, S460L, IT463-464VA, D470E, A482Q, KNV490-491-492TKT) which were related to strains detected in patients with gastric atrophy, gastric ulcer, and duodenitis. These mutations were also detected in Japanese and American strains isolated from patients with atrophy (Oki120, Oki112, Oki128, and Ok139 strains) ^(35, 36)^ and duodenal ulcer (Oki898 and Ok204 and J99 strains). ^(35-37)^ However, there are some strains of *H. pylori* implicated in both diseases gastric and duodenal ulcer (e.g., Germany strain UH44);^(38)^ and maybe Sudanese strains are one of them. Two strains (Sudan-1 and Sudan-3) were detected from patients with normal gastroduodenal finding had the same mutations of gastric ulcer strains. While the third one (Sudan-2) and gastritis strains share the same mutations of cancerous strain (A439P, T457P, N468K, and H500Y).The infection of those patients could be a new type and need more time to reflect the complications or could be due to host/environmental factors that delay the appearance of complications. ^(39-41)^ This finding is partially in agreement with the result obtained by Masahiko *et al*., that the Amino acid frequency of gastritis samples seems to be intermediate between gastric cancer and MALT lymphoma samples. ^(42)^ In contrast, two Japanese strains (NCTC11637 and NCTC11638) isolated from patients with gastritis do not have any mutations in this region (amino acid residues 428-510) except N467G which was found in all our sequences in regardless to gastroduodenal diseases.^(43)^

In this study, the frequency of *H. pylori* infection among endoscopics was found to be 37/50 (74%) using PCR; this result was higher compared to the prevalence of 18/81 (22.2%) reported by Mona *et al*., 2015 had been used culture methods. ^(44)^ Recently, assays based on PCR have been used to detect *H. pylori* DNA and its virulence genes in gastric biopsies with high specificity and quick results have been obtained without the requirement for special transport conditions. However, the presence of the *cagA* gene has been confirmed in 54% of the Sudanese *H. pylori* strains. The frequency of *cagA* positive *H. pylori* varies from one geographic region to another. In Sri Lanka is 48%, in the Netherlands is 67%, in Germany is 72%, in the United States is 81%, in Nigeria is 93% and in Korea is 97%. ^(2)^

The association between the genotype of *cagA* and clinical outcome was examined. There was no significant association between the genotype of *cagA* and clinical outcome (X^2^=1.567^a^; df=1; p=0.225). This finding agrees with the study conducted by Wen *et*.*al, 2004* in East Asia,^(45)^ and disagrees with a study that was done in Chinese population that showing 95% positive for *cagA gene* among patients with peptic ulcer disease and 100% with chronic gastritis.^(46)^This is could be related to geographic strain distribution ;therefore more studies should be considered to include more sample size include

## Conclusion

Disease-specific variation of *CagA* of *H. pylori* strains, in the region of amino acid residues 428-510, are found among Sudanese patients. A novel mutation (K458N) was detected in the basic amino-acid cluster, which provides a positive electrostatic surface for *CagA*. Phylogenetic analysis showed a high level of diversity of *cagA* among Sudanese *H. pylori* strains.

## Data Availability

All important data are available in the manuscript

